# Incidence and factors associated with infections in kidney transplant recipients at the tertiary hospital, Thailand

**DOI:** 10.64898/2026.09.28.26364156

**Authors:** Saowarat Phacharoen, Sukrit Kanchanasurakit, Korntip Phonphok, Duangjai Duangrithi, Papatsara Petchcharee, Wiriya Sangsanit, Rungrueng Jintasataporn, Abhisit Prawang

## Abstract

In medical practice, the rising prevalence of infections in kidney transplant recipients (KTRs) is a notable concern. However, understanding the incidence and risk factors associated with infections in KTRs remains limited. This study aims to assess the incidence and factors associated infection among KTRs. Additionally, we also investigate the median time of each infection among KTRs.

A retrospective cohort study was conducted from 1 January 2018-31 December 2022 at Thailand’s tertiary hospital. We performed univariable and multivariable logistic regression analyses to identify potential risk factors related infection and investigated for the incidence and median time of each infection among KTRs.

A total of 179 participants were included in this study. Our findings reveal the kidney from living donor was protective factor for infection among KTRs (aOR 0.063, 95% CI 0.005-0.792; *p* = 0.032). Urinary tract infections (UTIs)showed as the predominant pathogenic manifestation among KTRs (43%). The median time of UTIs, cytomegalovirus (CMV) infection and pneumocystis pneumonia (PCP) were 16, 57 and 94.5 days, respectively.

Our study establishes that receiving a kidney from a living donor is a protective factor against post-kidney transplant infections. Particularly urinary tract infections were most common of infection. Monitoring urinary tract infections post-transplantation is recommended. Moreover, awareness of UTI, CMV infection and PCP should be concerned during 2-3 months after transplantation. However, due to a small sample size; we recommend to increase the number of particular participants in further investigations.

## Introduction

Kidney transplantation remains the preferred strategy for end-stage kidney disease (ESKD), revealing superior survival and quality of life compared with long-term dialysis [1, 2]. However, infectious complications continue to be a major cause of morbidity and mortality among kidney transplant recipients (KTRs) [1, 3]. KTRs are needed for sustained immunosuppressive therapy for preventing rejection, substantially increases susceptibility to opportunistic and non-opportunistic infections. Balancing adequate level of immunosuppression with infection risk becomes a critical challenge in transplant pharmacotherapy.

In Thailand, patterns of post-transplant infections may differ from those reported in high-income settings due to variations in pathogen epidemiology, antimicrobial stewardship practices, vaccination administration policy, healthcare policy, and the way access to immunosuppressive agents. At Rajavithi hospital, a tertiary care hospital, where complex transplant cases are performed and managed, understanding infection incidence and potential risk factors is essential to optimize pharmaceutical care and infection prevention strategies regarding immunosuppressive regimens and antimicrobial utilization.

Previous studies have identified many factors associated with post-transplant infections, including type of immunosuppressive therapy, acute rejection episodes, comorbidities, donor characteristics, and delayed graft function. However, incidence of infection, time-to-event data remain limited in Southeast Asian settings [3, 4].

A cumulative infection is particularly relevant in transplant research because infection risk is dynamic and varies across post-transplant periods. Early infections are often related to surgical and healthcare-associated exposures, whereas late infections are more associated with long-term immunosuppression and community-acquired pathogens [1, 3, 4]. Categorizing infections by type, frequency of events, and timing allows for a more precise assessment of risk strategies and might inform policy decisions regarding type of transplantations, prophylactic strategies, immunosuppressive dose adjustment, and monitoring protocols.

In Thailand, data on infection patterns and survival outcomes among KTRs in tertiary-care settings remain limited. Generating locally relevant evidence is essential for informing national medical policy, including antimicrobial stewardship programs, prophylaxis guidelines, and cost-effective analysis of immunosuppressive therapies.

Therefore, this study aims to (1) identify factors associated with post-transplant infections among KTRs at a tertiary hospital in Thailand; (2) investigate incidence and median time of each infection among KTRs.

## Methods

### Study design and setting

A retrospective cohort study was conducted at Rajavithi hospital, a tertiary care hospital, in Thailand. The study included KTRs who underwent transplantation between 1 January 2018 - 31 December 2022.

### Study population

All adult patients (aged ≥18 years) who received a kidney transplant during the study period and had at least one follow-up visit after transplantation were eligible for inclusion. Patients with incomplete medical records or those who underwent multi-organ transplantation were excluded. Participants were followed from the date of transplantation until the occurrence of infection, graft failure, death, loss to follow-up, or the end of the study period (1 January 2018 - 31 December 2022), whichever occurred first. The size of population was calculated from Wayne WD, 1995 formulation [5]. When n was the required sample size (n = 172).

### Data collection

Clinical and pharmaceutical data were extracted from electronic medical records and medical sheets at transplant center of hospital. Variables collected included demographic characteristics (age, sex), comorbidities, donor type (living or deceased donor), induction therapy, maintenance immunosuppressive regimens, drug level, episodes of rejection, and mortality.

Infectious events were identified based on physician diagnosis supported by microbiological, radiological, or laboratory evidence, as documented in medical records. Infections were categorized by type, number of infectious episodes per patient, and timing relative to transplantation.

### Outcomes

The primary outcome was the risk factors associated infections among KTRs. Secondary outcomes included infectious events were categorized according to: type and number of infection and time to first infection event (reported as median time).

Time-to-event was defined as the duration (in days) from the date of kidney transplantation to the first documented infection.

### Statistical analysis

Descriptive statistics were used to describe baseline characteristics and infection patterns. Categorical variables were presented as frequencies and percentages, while continuous variables were reported as mean ± standard deviation or median with interquartile range, as appropriate. Univariate and multivariate logistic regression analysis was conducted to identify factors associated with time to first infection. Variables with p < 0.20 in univariate analysis were included in the multivariate model. Adjusted odd ratios (aOR) with 95% confidence intervals (CIs) were reported. A two-sided p-value < 0.05 was considered statistically significant. Statistical analyses were performed using standard statistical software.

### Ethical considerations

The study protocol was reviewed and approved by the ethics committee of Rajavithi hospital (No. 66059).

## Results

During the study period (2018–2022), a total of 179 patients underwent kidney transplantation at the tertiary hospital met the inclusion criteria and were included in our analysis. In our infection study group (Table 1.); The median age of participants was 42.5 years with an interquartile range (IQR) 37.00-50.25 years, and 56.20% were male. The majority of patients received kidneys from 15 living donors (12.8%). The most cause, underlying disease and renal graft function of KT was unknown (48.4%), hypertension (38.2%) and immediate/slow (68.3%), respectively. Interestingly, the rate of acute rejection and survival are desirable which are reported as 3.4% and 89.2%, respectively.

**Table 1.** Demographic and clinical characteristics in kidney transplant recipients at the tertiary hospital, Thailand recipients at the tertiary hospital, Thailand.

| Variables | Infection |  |
| --- | --- | --- |
|  | No<br>n (%) | Yes<br>n (%) |
| Male | 21 (37.50) | 68 (56.20) |
| Type of KT; LD | 14 (25.50) | 15 (12.80) |
| Cause of KT |  |  |
| Unknown | 30 (53.60) | 59 (48.40) |
| Presume GN | 4 (7.10) | 7 (5.70) |
| Polycystic kidney disease | 0 | 8 (6.60) |
| Obstructive nephropathy | 1 (1.80) | 2 (1.60) |
| LN | 1 (1.80) | 4 (3.30) |
| DN | 6 (10.70) | 12 (9.80) |
| NSAIDs | 1 (1.80) | 5 (4.10) |
| Chronic glomerulonephritis | 12 (21.40) | 15 (12.30) |
| Kidney injury gunshot wound | 0 | 1 (0.80) |
| Renal from hypertension | 1 (1.80) | 9 (7.40) |
| ABO blood group |  |  |
| A | 13 (24.10) | 16 (13.10) |
| A+ | 1 (1.90) | 1 (0.80) |
| AB | 5 (9.30) | 10 (8.20) |
| AB+ | 1 (1.90) | 0 |
| B | 17 (31.50) | 49 (40.20) |
| B+ | 0 | 2 (1.60) |
| O | 17 (31.50) | 44 (36.10) |
| Underlying diseases |  |  |
| No | 7 (12.50) | 12 (9.80) |
| HT | 21 (37.50) | 47 (38.20) |
| DLP | 0 | 2 (1.60) |
| MI | 0 | 1 (0.80) |
| Hyperparathyroidism | 1 (1.80) | 0 |
| Thalassemia | 0 | 1 (0.80) |
| SLE | 1 (1.80) | 0 |
| HBs Ag: positive | 1 (1.80) | 4 (3.30) |
| HCV: positive | 1 (1.80) | 1 (0.80) |
| Sex of donor: male | 29 (76.30) | 62 (77.50) |
| Prolong cold ischemia >24 h | 8 (14.30) | 19 (15.80) |
| HLA mismatch: point score 0-3 | 40 (72.70) | 89 (74.20) |
| ABO incompatibility | 2 (3.60) | 2 (1.70) |
| Previous rejection | 4 (7.10) | 7 (5.80) |
| Tacrolimus subtherapeutic level | 45 (95.70) | 90 (86.50) |
| Tacrolimus suprathapeutic level | 16 (40.00) | 39 (40.20) |
| Cyclosporine-included regimen | 12 (22.20) | 20 (16.70) |
| Renal graft function; immediate or slow | 41 (80.40) | 82 (68.30) |
| Acute rejection | 8 (14.50) | 16 (13.40) |
| Chronic rejection | 6 (11.30) | 17 (14.30) |
| Survive | 55 (98.20) | 107 (89.20) |
| Prolong HD >5 years | 16 (30.20) | 60 (52.60) |
| Prolong hospitalization >30 days | 20 (36.40) | 65 (56.50) |
| Prior VZV infection | 1 (1.80) | 8 (6.50) |
| CMV D+/R- | 1 (1.8) | 8 (6.5) |
| Scr >10 mg/dL before KT | 26 (46.40) | 48 (39.30) |
| Scr at discharge <1.5 mg/dL | 34 (60.70) | 69 (56.60) |
| Transplant duration (years) (median; IQR) | 8.00 (6.00-10.00) | 7.00 (5.00-10.00) |
| Age (years) (median; IQR) | 44 (32.25-50.50) | 42.50 (37.00-50.25) |
| Age of donor (years) (median; IQR) | 40.00 (27.25-52.00) | 41.50 (31.75-53.25) |

CVD; cardiovascular diseases; CMV; cytomegalovirus; DLP; dyslipidemia; DN: Diabetes nephropathy; DM: Diabetes mellitus; GN: glomerulonephritis; HBs Ag: Hepatitis B surface antigen; HCV: hepatitis C virus; HD: hemodialysis; HLA: human leukocyte antigen; HT: hypertension; IQR: Interquartile Range; KT: kidney transplant; LD: living donor; LN: lupus nephritis; MI: myocardial infarction; NSAIDs: nonsteroidal anti-inflammatory drugs; SLE; Systemic lupus erythematosus; Scr: serum creatinine; VZV: varicella Zoster Virus

On univariate analysis, we analyzed these factors: sex, type of KT, HBs Ag: positive, HCV: positive, sex of donor, prolong cold ischemic time, HLA mismatch, ABO incompatibility, previous rejection, tacrolimus supratherapeutic level, cyclosporine-included regimen, renal graft function, acute rejection, chronic rejection, prolong HD, prolong hospitalization, prior VZV infection, CMV D+/R-, transplant duration, age of KT recipient and age of donor. The factors with p-value < 0.2 have been analyzed on multivariate analysis combined with type of KT, renal graft function, prolong HD, prolong hospitalization and transplant duration. On multivariate analysis, the only factor significantly (adjusted OR = 0.063; 95% CI: 0.005-0.792; p = 0.032) associated with infections was Type of kidney transplant: living donor (Table 2).

**Table 2.** Multivariate ORs (95% CI) of predicting factors associated with infections in kidney transplant recipients at the tertiary hospital, Thailand.

| Factor | Multivariate* |  |
| --- | --- | --- |
|  | aOR (95% CI) | P |
| Type of kidney transplant: living donor | 0.063 (0.005-0.792) | 0.032 |

\*Adjusted for age, gender, sex of donor, hepatitis B and C infection, acute and chronic rejection.

### Incidence and characteristics of infectious events

Overall, 123 patients (68.72%) developed at least one post-transplant infection during follow-up. At day 30 post-transplant revealed top 3 of infections were urinary tract infection (UTI) (43%), other infections (nocardiosis, tuberculosis, septicemia, Chicken pox, aspergillosis, Dengue fever, *C. difficile* infection and intra-abdominal infection) and HPV infection. At day 30-180 post-transplant revealed top 3 of infections were urinary tract infection, other infections and CMV infection. At day > 180 post-transplant revealed top 3 of infections were the same of day 30-180 (Figure 1). The median time of UTIs, cytomegalovirus (CMV) infection and pneumocystis pneumonia (PCP) were 16, 57 and 94.5 days, respectively. The median time of each infection was shown in Table 3.

**Table 3.**
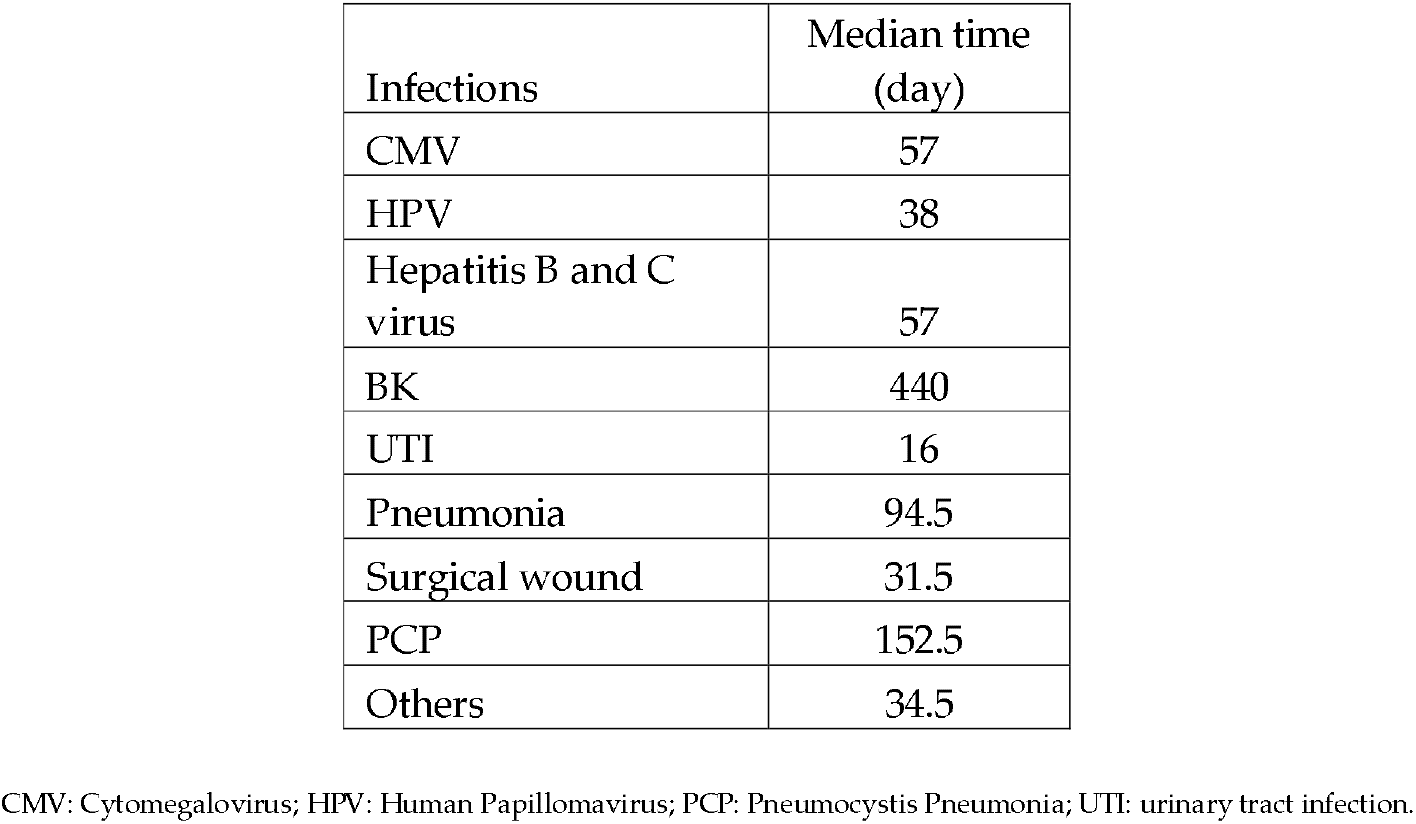
The median time of infections in kidney transplant recipients.

| Infections | Median time (day) |
| --- | --- |
| CMV | 57 |
| HPV | 38 |
| Hepatitis B and C virus | 57 |
| BK | 440 |
| UTI | 16 |
| Pneumonia | 94.5 |
| Surgical wound | 31.5 |
| PCP | 152.5 |
| Others | 34.5 |

CMV: Cytomegalovirus; HPV: Human Papillomavirus; PCP: Pneumocystis Pneumonia; UTI: urinary tract infection.

**Figure 1.**
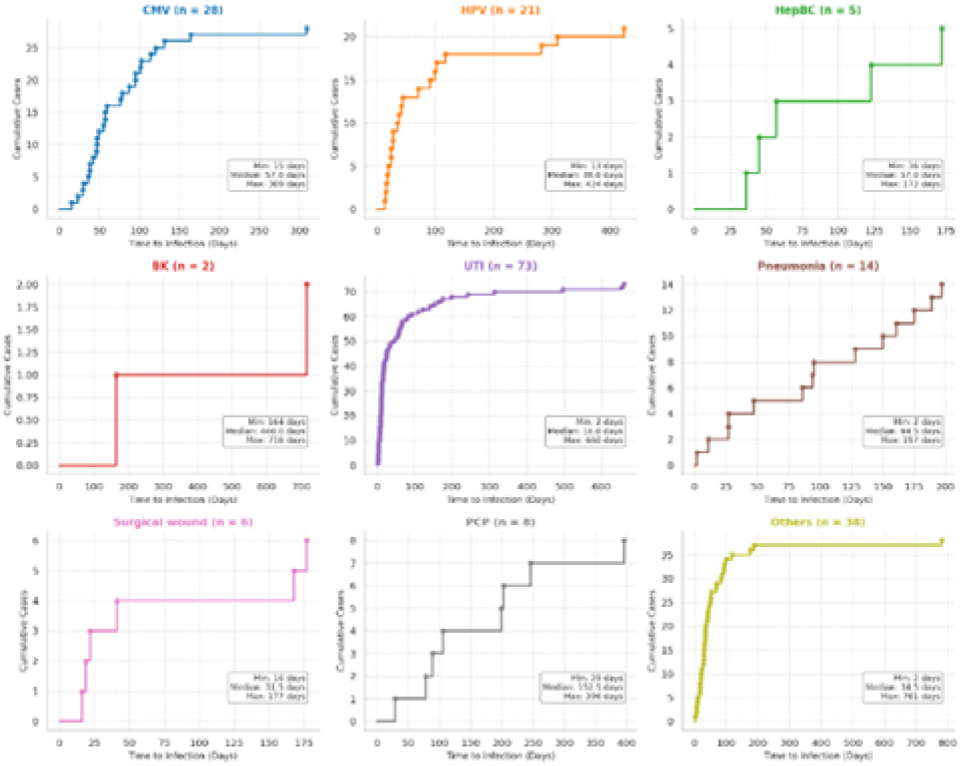
Cumulative infections in kidney transplant recipients.

## Discussion

In this retrospective study of KTRs at a tertiary care hospital, post-transplant infections were high, affecting approximately two-thirds of patients during the follow-up period. Despite the burden of infectious complications, patient survival remained favorable, with an overall survival rate of 89.2% and a low incidence of acute rejection. These findings highlight the continuing challenge of infection management in KTRs while demonstrating the effectiveness of contemporary immunosuppressive and post-transplant care strategies.

The overall infection rate of 68.7% observed in our study is consistent with previous reports indicating that infections remain one of the leading causes of morbidity among KTRs [1, 3, 4]. The high infection rate in this population is attributable to chronic immunosuppression, prolong hospitalization, and the presence of multiple comorbidities.

The infections were observed in this study generally followed the well-established pattern of post-transplant infectious syndromes. UTI was the most common early infection, with a median onset of 16 days after transplantation. This finding is expected because urinary catheterization, ureteral stent placement, perioperative manipulation of the urinary tract, and intensive immunosuppression during the early post-transplant period increase susceptibility to bacterial infections. Previous studies have similarly identified UTI as the most frequent infectious complication among KTRs, particularly during the first month after transplantation [6, 7].

Viral infections, Cytomegalovirus (CMV) infection occurred at a median of 57 days post-transplantation, corresponding to the period of maximal immunosuppressive exposure. CMV remains one of the most clinically significant opportunistic infections after kidney transplantation because of its direct effects on organ systems and its indirect effects on graft function and immune regulation. The timing observed in our study is comparable to previous reports, which typically describe CMV disease occurring within the first three to six months following transplantation, particularly among high-risk recipients [3, 4].

Interestingly, BK virus infection occurred substantially later than other infectious complications, with a median onset of 440 days after transplantation. This finding is consistent with the pathogenesis of BK virus nephropathy, which often develops after prolonged immunosuppressive exposure [8]. The delayed occurrence emphasizes the importance of long-term surveillance beyond the first post-transplant year, particularly in patients receiving maintenance immunosuppressive regimens.

PCP was observed at a median of 152.5 days after transplantation, while pneumonia from other causes occurred at a median of 94.5 days. These findings support the vulnerability of transplant recipients to opportunistic respiratory infections during the intermediate post-transplant period [9]. This finding may suggest antibiotic prophylaxis such as co-trimoxazole among KTRs.

Our analysis identified the type of kidney transplantation as the only factor associated with post-transplant infection in the multivariable model. Patients receiving living-donor kidney transplants appeared to have a lower risk of infectious complications compared with recipients of deceased donor. Although the confidence interval was relatively wide, this association may reflect several advantages of living-donor transplantation, including shorter ischemic times, better graft quality, earlier graft function recovery, shorter hospitalization and fewer donor-derived complications. Previous studies have reported superior graft outcomes and reduced complication rates among living-donor transplant recipients, which may contribute indirectly to a lower infection risk [10, 11, 12]. We highly recommend to undergo living-donor kidney transplant if there is possibility.

In contrast, other potential risk factors, including recipient age, donor age, hepatitis B or C status, HLA mismatch, acute rejection, prolonged hospitalization, and supratherapeutic tacrolimus concentrations, were not associated with infection after adjustment. These findings may be attributable to the relatively limited sample size, low prevalence of some risk factors, or effective clinical management practices that affect their impact.

However, the study also has limitations. Its retrospective design may have resulted in incomplete data collection and potential misclassification of infectious events. The single-center setting may limit the generalizability of the findings to other transplant populations. Future multicenter studies with larger sample sizes and longer follow-up periods are needed to validate these findings and further characterize specific risk factors for mortality [13, 14].

## Conclusion

In conclusion, infectious complications remain common after kidney transplantation, affecting more than two-thirds of recipients in this study. UTI was the predominant early infection, CMV was the most common viral infection during the intermediate post-transplant period, and BK virus infection occurred predominantly during long-term follow-up. Living-donor kidney transplantation was associated with a lower risk of post-transplant infection. These findings provide valuable real-world evidence regarding the incidence and timing of post-transplant infections and may help optimize infection prevention, monitoring, and management strategies in kidney transplant recipients.

## Data Availability

All data produced the present work are contained in the manuscript

## Supplementary Materials

## Author Contributions

Conceptualization, S.P., A.P., D.D., K,P. and S.K.; Data curation, P.P., W.S., and R.J.; Formal analysis, A.P., S.K., D.D., and K.P.; Investigation, A.P., S.P., and S.K.; methodology, S.P., A.P., and D.D.; Writing-original draft: S.P., A.P., and D.D.; Writing-review & editing, S.P., D.D., and S.K. All authors have read and agreed to the published version of the manuscript.

## Funding

This research received no external funding.

## Informed Consent Statement

Patient consent was waived due to the participants would not be exposed to an increased risk of harm.

## Data Availability Statement

## Acknowledgments

We would like to thank the Kidney transplant team at Rajavjthi hospital for data base support.

## Conflicts of Interest

The authors declare no conflict of interest.

## Notes

### Competing Interest Statement

The authors have declared no competing interest.

### Author Declarations

The study protocol was reviewed and approved by the ethics committee of Rajavithi hospital (No.66059).

